# Hidden Map of the Human Cochlea and the Law of Cochlear Degeneracy

**DOI:** 10.64898/2026.11.24.26368840

**Authors:** Zakarya Hadj-Allal, Antti Aarnisalo, Antti A. Mäkitie

## Abstract

For a century, the pure-tone audiogram has been the foundation of hearing science and otology, routinely assumed to reflect specific cochlear pathology. Here, we resolve the mapping from macroscopic auditory function to microscopic pathology by matching 4.2 million human audiograms against 37 million cochlear states generated using four cochlear model simulators. We show that the audiogram does not specify a unique underlying tissue state: in at least 97.3% of ears, the same audiogram was produced by distinct categories of cochlear cellular lesions. This ambiguity follows two rules, which we formulate as the Law of Cochlear Degeneracy. First, outer and inner hair-cell injuries replace one another only at the same place along the cochlea. Second, only up to a limit: this mutual substitution holds across mild loss but collapses as impairment deepens, and in the tested models outer hair-cell injury alone reaches no more than 44 dB on average. Because the cochlea cannot be biopsied during life, we built a population-scale library of cochlear lesions we call the Hidden Map of the Cochlea, which lists for every audiogram the cellular lesions compatible with it. For any audiogram, the map then names the physiological measurements that tell those lesions apart, a step we call the Cochlear State Differential. Where audiograms are identical, otoacoustic emissions and envelope-following responses separate the underlying cochlear lesions, isolating mechanical amplification loss from sensory transduction injury. Reading the cochlea from the audiogram thus becomes a computable biophysical differential.

**Significance Statement:** For a century, hearing loss has been diagnosed by audiogram, assumed to reveal which cochlear cells are damaged. Because the living cochlea cannot be biopsied, that assumption was never tested at population scale. Matching 4.2 million human audiograms against 37 million cochlear states simulated in four cochlear models, we show it fails in nearly every ear: at least 97.3% are consistent with more than one type of hair-cell lesion. Temporal-bone studies showed such ambiguity exists; we establish the rules governing it: which injuries mask one another, where, and up to what severity. As gene and cell therapies target specific cochlear cells, selecting patients by audiogram risks treating different diseases as one. Our results specify noninvasive biomarkers predicted to decouple them.

## Introduction

Hearing loss affects more than 1.5 billion people worldwide and is the third leading cause of years lived with disability (1, 2). The World Health Organization projects that by 2050 nearly 2.5 billion people will be living with some degree of hearing loss, and estimates that unaddressed hearing loss already costs the world almost US$1 trillion each year (2). For more than a century its diagnosis has centred on the pure-tone audiogram, standardised as a clinical classification tool in the 1940s (3) and read, from the classic audiometric and histological schemes onward, as reflecting specific cochlear lesions (4). Whether those audiograms also carry a reproducible physiological state, and what that state does and does not specify about the cochlea, has not been established.

We previously showed that human hearing does not come in categories but occupies a continuous five-dimensional geometric space transferable across populations, eras and calibration standards, which we called the Human Hearing Atlas, and that five Hearing Atlas Coordinates locate any audiogram and reconstruct unseen audiograms to near-decibel accuracy (5). The Atlas does not by itself identify the anatomical or molecular cause of a given hearing state, leaving the relationship between cochlear states and hearing states open to investigation. Here, we link cochlear states to the audiogram. Describing a patient’s hearing state does not reveal the underlying tissue damage. The cochlea is encased in the densest bone of the body and cannot be biopsied during life, so the inference from audiogram to tissue lesion is made blind. The audiogram contains the information on a hearing state and always has; it cannot independently distinguish whether the specific cells producing it are intact, damaged, or completely missing.

A century of temporal-bone histopathology has nevertheless established that similar audiograms can accompany markedly different cochlear pathology (6, 7, 8, 9), and that substantial neural injury can occur despite a preserved audiogram (10, 11). Conversely, changing inner hair cell (IHC) synapse density leaves brainstem and otoacoustic sensitivity unchanged while neural amplitude and temporal processing move from impaired to supranormal (12). These observations remain descriptive: they show that audiograms can fail to identify tissue injury, but not whether this ambiguity follows reproducible rules.

Here we overturn that reading and establish that the cochlea is physiologically degenerate (13), meaning that entirely distinct cellular states can produce the exact same observable audiogram (14, 15, 16). The questions then become which injuries are interchangeable, whether that interchangeability follows quantitative rules, how it changes as hearing deteriorates, and what additional physiology reveals differences concealed by the audiogram.

We therefore mapped every hearing state back to the cochlear states that produce it (17, 18, 19, 20). Rather than fitting one cochlear state to each audiogram, we generated audiograms from specified cochlear states and retained every modelled state that reproduced a patient’s measured audiogram. This global reverse-lookup generates a population-scale library of cochlear lesions we call the Hidden Map of the Cochlea Library. When applied to an individual patient, this lookup yields their Hearing-State Cochlear Map, identifying the full set of cochlear lesions compatible with their audiogram. To resolve this ambiguity requires a Cochlear State Differential: a targeted secondary physiological exam, which narrows the compatible cochlear lesions to yield the final Differential Cochlear States. Finally, comparing these equivalent states across the population shows that outer hair cell (OHC) and IHC injuries replace one another only at the same cochlear place, and only up to a limited severity. We call these rules the Law of Cochlear Degeneracy.

## Results

### A reverse map from the audiogram to every compatible cochlear state

#### Four cochlear descriptions reproduced different fractions of the same audiograms

Of 4,210,288 six-frequency audiograms, combined OHC and IHC dysfunction (BEZ2018/Zilany) reproduced 99.7% within 5 dB at every frequency (4,197,616), cochlear frequency-tuning mechanics (CARFAC v2) 67.5% (2,843,632), OHC and basilar-membrane mechanics alone (Verhulst 2018) 66.5% (2,798,252) and a phenomenological model of five cochlear and middle-ear mechanisms (M0) 42.7% (1,796,448). The disagreement lay inside individual audiograms: the two OHC models each reproduced about one audiogram in nine that the other did not (503,032 for CARFAC v2 and 457,652 for Verhulst 2018), and BEZ2018/Zilany reproduced every audiogram that either of them did, together with 32.2% and 33.2% that CARFAC v2 and Verhulst 2018 did not (**SI Appendix, Fig. S1a**). That difference establishes broader model reachability, not IHC involvement in those ears.

#### We built a Hidden Map of the Cochlea Library totalling 37,137,415 simulated cochlear states, together with the six-frequency hearing state each one generates

The library spans four cochlear models: 29 million combined OHC and IHC states (sampling the global joint domain alongside targeted isolated-injury and near-normal regimes, with targeted enrichment in sparse regions of the hearing-state space), 4 million states restricted to OHC and basilar-membrane mechanics, 3 million states of the five-mechanism model (M0), and 1 million states of cochlear amplification and frequency tuning. The isolated-injury and near-normal states test whether near-normal, OHC-only, or IHC-only tissue categories remain compatible with a given hearing state, while the global states sample the joint OHC to IHC domain. Every state retained its physiological parameters and its generated audiogram and was indexed by the same five Atlas coordinates used to describe the patients. Matching an audiogram against the library returns every compatible cochlear state rather than one selected reconstruction, and retrieval was verified against exhaustive candidate examination.

#### The same audiogram can be produced by different cochlear abnormalities; the audiogram alone therefore does not uniquely identify the underlying lesion

At the 5 dB step of clinical audiometry (21), at least 97.3% of 4,210,288 six-frequency audiograms were compatible with more than one lesion category, 2.4% with exactly one and 0.3% with none in the library, from 88.8% to 97.6% across the four contributing surveys (**Fig. 1; SI Appendix, Figs. S1b and S3 to S8**). The multi-category lower bound was 97.3%, 97.3% and 97.3% with impairment defined below 0.98, 0.90 and 0.80 of the healthy value (SI Appendix, Table S3), and 89.1% among the 959,027 ears with a mean hearing level of 20 dB or more. Among the 2,798,252 audiograms that an OHC and basilar-membrane model could reproduce, every one also admitted a mixed or IHC alternative.

**Fig. 1.**
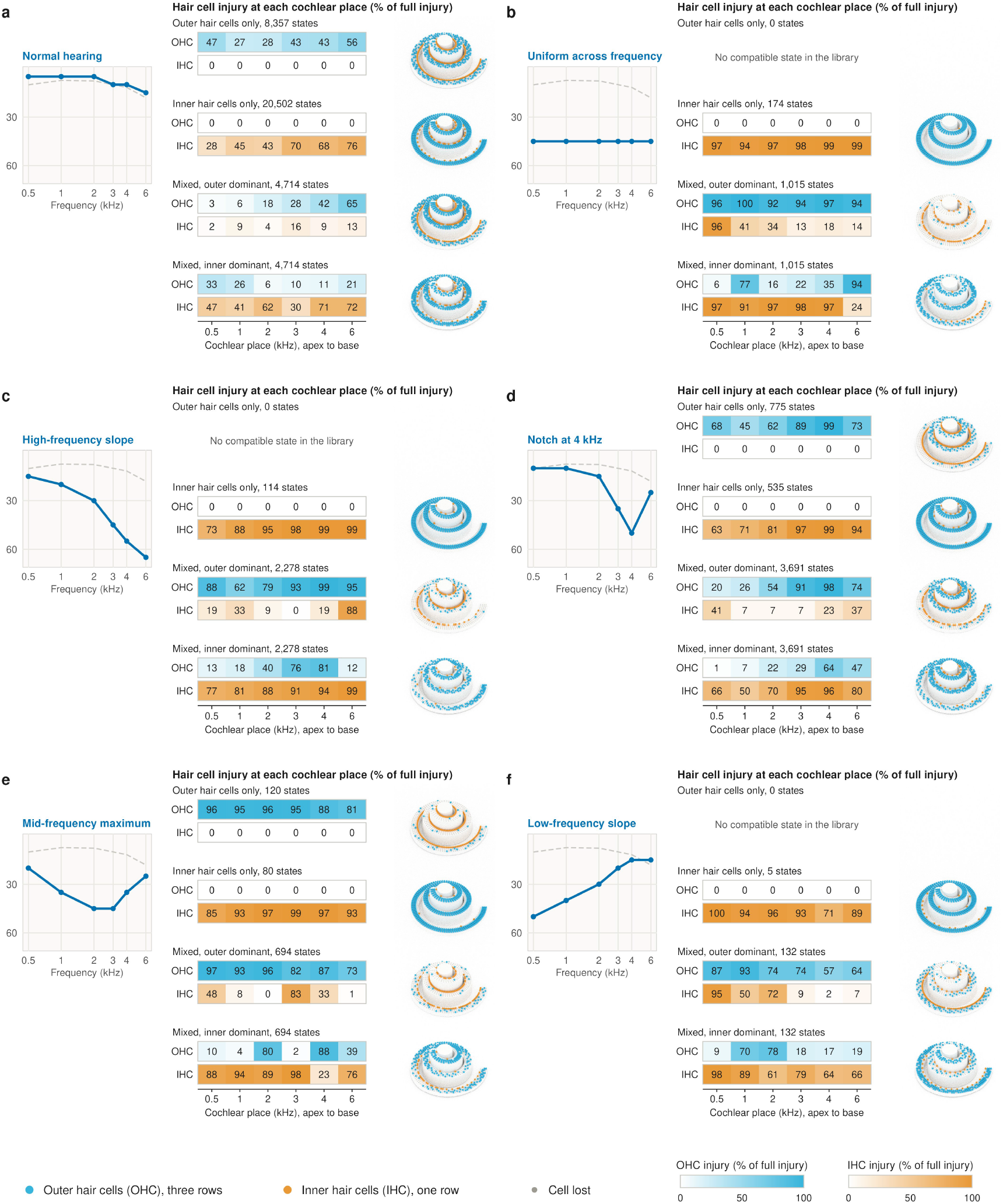
Hair-cell injury profiles of simulated cochlear states that reproduce six reference audiograms, by lesion category. a-f, Six reference audiograms. In each panel: left, the six-frequency audiogram (hearing state; solid line) against the population median (dashed line), hearing level increasing downward with ticks at 30 and 60 dB HL; centre, simulated cochlear states (BEZ2018) from its Hearing-State Cochlear Map, each reproducing the audiogram within 5 dB at every frequency. Each number is the injury of outer hair cells (OHC) or inner hair cells (IHC) at that cochlear place, as a percentage of full injury (0, intact; 100, no remaining function of that cell type in the model); each pair of rows is headed by the lesion category and the number of library states of that category that reproduce the audiogram; right, the same states drawn as cochleae from the base (6 kHz, outer end) to the apex (0.5 kHz, centre), where the proportion of cells drawn as lost at each place equals the fractional injury. Where no library state of a lesion category reproduces the audiogram, no state is drawn. HL, hearing level.

#### OHC and basilar-membrane dysfunction alone cannot explain the most severe audiograms within the tested cochlear models

Verhulst 2018 generated no six-frequency mean hearing level above 36.98 dB and CARFAC v2 none above 33.0 dB. Of 161,053 audiograms with a mean severity of 40 dB or more, CARFAC v2 reproduced none and Verhulst 2018 seven. Above the hearing levels an isolated OHC model can generate, a broader Hearing-State Cochlear Map is required.

#### Only 0.24% of ears (10,270/4,210,288) lay outside all four models

from 0.06% in the occupational archive to 10.7% in the 1963 to 1970 survey; a further 2,402 ears outside the BEZ2018/Zilany bank were reproduced by the five-mechanism model (M0) alone. These ears set a population benchmark: a future cochlear model that reproduces part of the region increases explanatory coverage of population audiograms, and one that does not leaves this coverage gap unchanged.

### Different cochlear injuries can produce the same hearing state

A reproducible hearing state does not uniquely specify the cochlear physiology that produced it. Across 4,210,288 six-frequency audiograms matched against the library, multiple distinct states remained compatible with the same audiogram: **physiological degeneracy**, in which different cochlear states are functionally indistinguishable on the audiogram.

Within the place-resolved OHC and IHC banks, most sampled OHC states had an IHC state reproducing the six-frequency audiogram inside the prespecified 5-dB criterion: 2,434,758 of 2,593,563 OHC queries matched, a support fraction of 0.939, whereas 76,551 of 99,545 IHC queries had an admissible OHC match, 0.769 (**Figs. 1 and 2**).

**Fig. 2.**
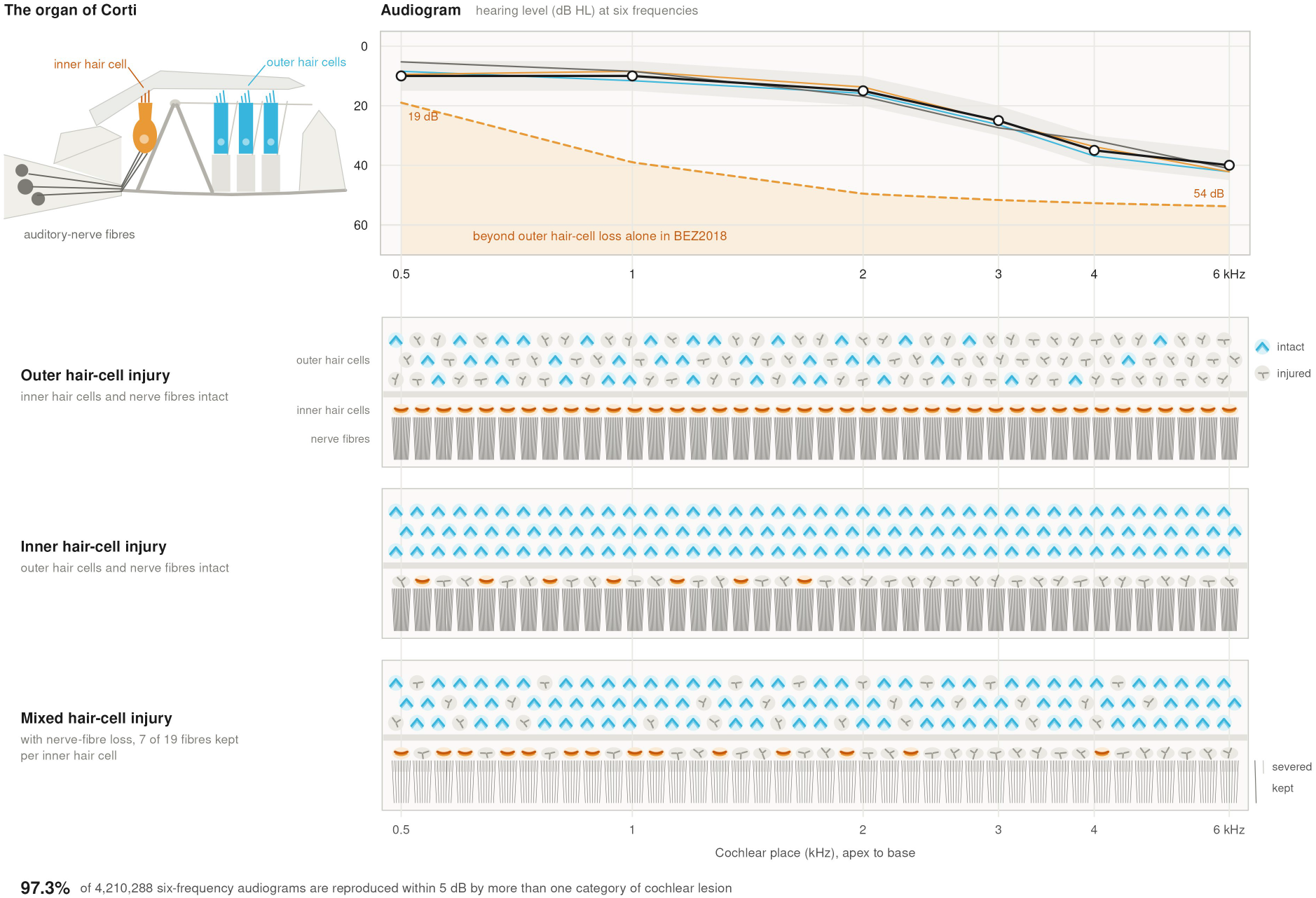
Outer hair-cell, inner hair-cell and combined injury states that reproduce the same measured audiogram. Top, one six-frequency audiogram from the census (black; band, 5 dB either side) and the audiograms generated by the three cochleae below it (thin lines, in their colours). Orange dashed line, the hearing level generated at each frequency by complete outer hair-cell loss with intact inner hair cells in BEZ2018 (19 dB at 0.5 kHz to 54 dB at 6 kHz; mean of the six, 44.3 dB); an audiogram in the shaded region is not produced by outer hair-cell injury alone in that model. Below, three cochleae that reproduce the audiogram within 5 dB at every frequency, each drawn as the organ of Corti seen from above, from the 0.5 kHz place (left) to the 6 kHz place (right), with three rows of outer hair-cell bundles (blue), one row of inner hair-cell bundles (orange) and the radial auditory-nerve fibres (dark grey), 19 per inner hair cell. A grey, splayed bundle marks a cell drawn as injured; the proportion so drawn at each place equals the fractional injury of that cell type there. The first two are BEZ2018 states with injury of outer hair cells only (53 to 94% from 0.5 to 6 kHz; 639 library states of this category reproduce the audiogram) and of inner hair cells only (63 to 98%; 518). The third is a BEZ2018 state with injury of both (outer 5 to 38%, inner 13 to 99%; 3,677 mixed states), drawn with loss of nerve fibres, 7 of 19 kept per inner hair cell, the more severe profile tested in Verhulst 2018. BEZ2018 has no fibre parameter; in Verhulst 2018 fibre loss leaves the modelled audiogram unchanged, because the hearing level is read from one high-spontaneous-rate fibre at the characteristic place. Fibres of the first two cochleae are drawn intact. Upper left, the organ in section, for orientation. Of 4,210,288 six-frequency audiograms, 97.3% were compatible with more than one lesion category of BEZ2018 states at the 5 dB criterion, a lower bound. Cell shapes carry no quantity. HL, hearing level.

We then tested hearing-state equivalence in a second cochlear model in which pole-dominant mechanical injury and *G*_met_-dominant transduction injury varied independently. Four prespecified targets with high-frequency slope from 15 to 30 dB were supported, forming a contiguous interval bounded below and above by unsupported targets and yielding downstream targets at 19.5, 24.0 and 28.5 dB with 23,045, 57,457 and 27,954 admissible pole and *G*_met_ pairs at the primary 5-dB criterion.

Equivalent flat hearing states were not found at any of ten flat targets, so hearing-state equivalence occupied a restricted part of the severity regime.

### OHC and IHC injuries substitute predominantly at corresponding cochlear places

We first quantified the strength of same-place substitution in BEZ2018, where frequency channels are independent by construction, and then tested spatial locality directly in Verhulst 2018, where places are coupled by the travelling wave. A model containing only OHC injury at the same cochlear place as the IHC response already predicted the matched IHC injury well: across 200 grouped train-test splits, held-out *R*^2^ ranged from 0.843 at 500 Hz to 0.900 at 1 kHz and stayed between 0.893 and 0.895 from 2 to 6 kHz, with median absolute errors of 0.071 to 0.078 in fractional-damage units (**Table 1**).

**Table 1.** Same-place substitution between outer and inner hair-cell injury, by frequency.

| **Frequency (Hz)** | **Same-place slope** | **Held-out *R*<sup>2</sup>, same place** | **Median absolute error** | **Held-out *R*<sup>2</sup>, full contextual model** |
|---|---|---|---|---|
| 500 | 0.7969 | 0.8429 | 0.0749 | 0.8460 |
| 1,000 | 0.9294 | 0.8996 | 0.0712 | 0.9114 |
| 2,000 | 0.9532 | 0.8948 | 0.0761 | 0.9022 |
| 3,000 | 0.9693 | 0.8938 | 0.0773 | 0.9066 |
| 4,000 | 0.9737 | 0.8939 | 0.0773 | 0.9081 |
| 6,000 | 0.9768 | 0.8930 | 0.0776 | 0.9051 |

*Slopes are the same-place coefficient of the model containing only outer hair-cell injury at the cochlear place of the inner hair-cell response. Held-out values are medians over 200 grouped train-test splits. Errors are in fractional-damage units.*

Same-place slopes were 0.797, 0.929, 0.953, 0.969, 0.974 and 0.977 from 500 Hz through 6 kHz, so from 1 to 6 kHz an increase in OHC injury at one place was accompanied by an almost proportional increase in the IHC injury required at that place to preserve the state-matched state. The relationship was weaker at 500 Hz but remained strongly predictive.

Adding injury at the other five places, neighbouring-place injury and seven prespecified topology descriptors increased held-out *R*^2^ by only 0.003 to 0.014. The dominant rule was local.

Of 10,400 attempts in Verhulst 2018 to offset a local loss of OHC gain by a change at another place, none of the 9,694 made a quarter octave or more away succeeded, and those made closer succeeded in 23.0% and 41.6% at the two baseline injuries tested (**Fig. 3a**). Substitution was therefore confined to about a quarter octave, the test resolving 0.076 octave. This bound concerns compensation of active mechanical gain along the travelling wave, not detection by off-place listening at high levels in complete IHC dead regions (22).

**Fig. 3.**
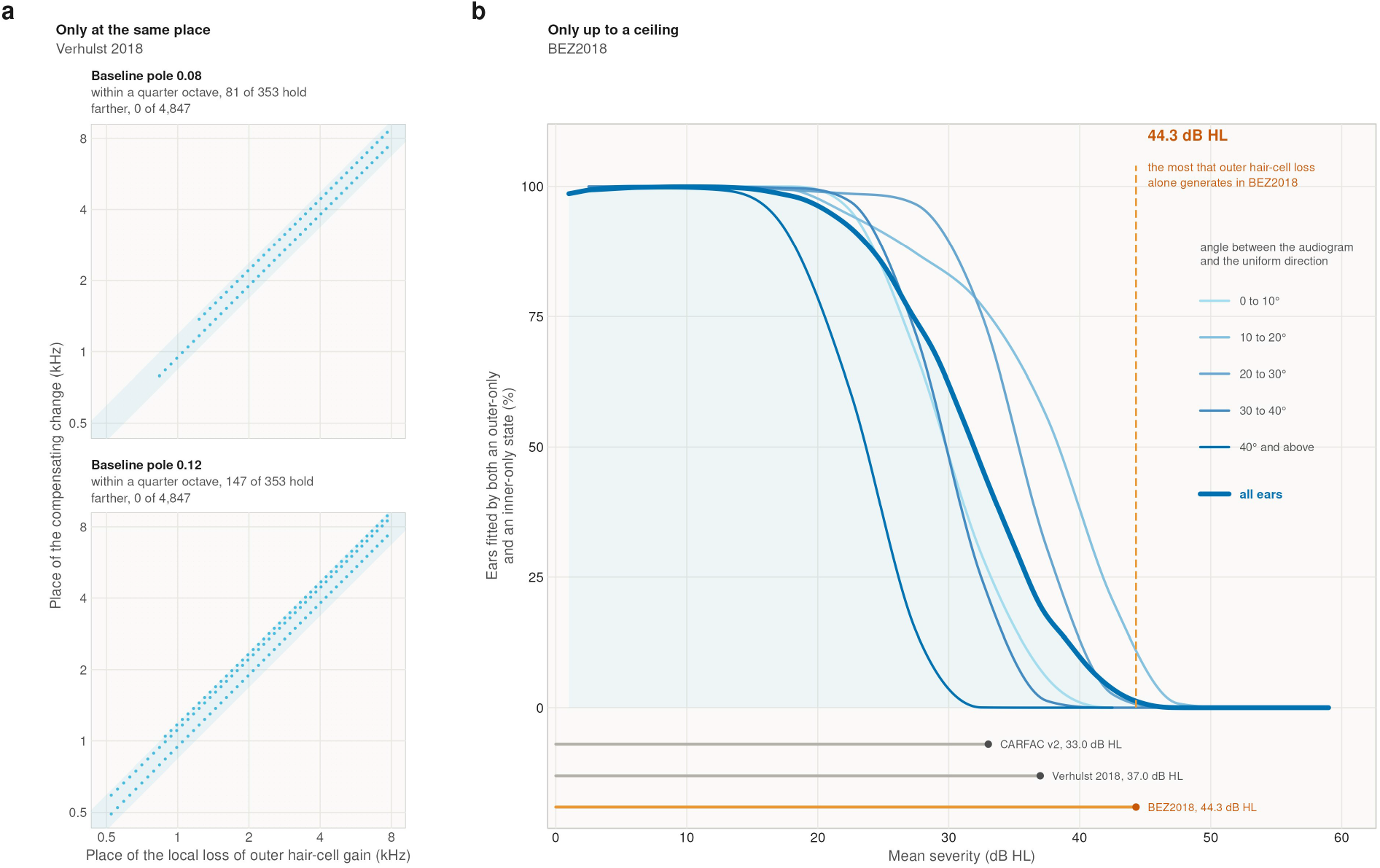
Spatial extent and severity limit of substitution between outer and inner hair-cell injury. (a) Only at the same place. In Verhulst 2018, where cochlear places are coupled, a loss of outer hair-cell gain over half an octave centred at one place was offset by a compensating change of gain centred at another, from two uniform baselines (pole 0.08 and 0.12; the normal-hearing value is 0.036). Blue, pairs in which the hearing level at the injured place returned within 2.5 dB of its baseline; faint grey, pairs in which it did not; shaded band, a quarter octave either side of the injured place. None of the 9,694 pairs centred a quarter octave or more away held. The window runs from 0.43 to 9.2 kHz on both axes; the 4,368 pairs outside it did not hold. Neighbouring centres are 0.071 to 0.088 octave apart, the resolution of the test. The test concerns compensation of active mechanical gain within one injury type, not detection by off-place listening in inner hair-cell dead regions. The same-place slopes of BEZ2018 are given in Table 1. (b) Only up to a ceiling. Share of ears reproduced within 5 dB by both an outer-hair-cell-only and an inner-hair-cell-only state of BEZ2018, against the mean of the six hearing levels. Dark line and shading, all ears in 2 dB bins (n = 4,194,057). Lighter lines, the same share in 5 dB bands by the angle between the six-frequency audiogram and the uniform direction (n = 4,168,949; cells with fewer than 200 ears are not drawn); curves pass smoothly through the tabled values. Orange dashed line, 44.3 dB HL, the mean hearing level generated by complete outer hair-cell loss with intact inner hair cells in BEZ2018; from 44 dB HL the share is at most 0.7%, and none from 48 dB HL. Bars beneath the axis, the largest mean hearing level that outer hair-cell injury alone generates in each model (CARFAC v2 33.0, Verhulst 2018 37.0, BEZ2018 44.3 dB HL). The decline does not exceed what a reach ceiling alone produces. HL, hearing level.

### Interchangeability ends at a ceiling, and the specific hearing state matters at higher severity

State-matched states did not remain interchangeable everywhere. In the simulated banks the outer-to-inner matched fraction held at 1.0 through seven burden deciles and fell to zero by the tenth (**Table 2**), and the reverse direction declined continuously with separation (**Table 3**).

**Table 2.** Outer-to-inner hair-cell support by injury burden.

| Burden decile | States queried | Matched | Support fraction |
|---|---|---|---|
| 1 | 109 | 109 | 1.0000 |
| 2 | 1,190 | 1,190 | 1.0000 |
| 3 | 1,441 | 1,441 | 1.0000 |
| 4 | 1,417 | 1,417 | 1.0000 |
| 5 | 1,442 | 1,442 | 1.0000 |
| 6 | 1,429 | 1,429 | 1.0000 |
| 7 | 1,442 | 1,442 | 1.0000 |
| 8 | 1,455 | 876 | 0.6021 |
| 9 | 1,162 | 24 | 0.0207 |
| 10 | 110 | 0 | 0.0000 |

*Flat injury category, outer-to-inner hair-cell direction, at the primary 5 dB state-matching criterion. Support is the fraction of queried states with an admissible counterpart in the other injury family.*

**Table 3.** Inner-to-outer hair-cell coverage by hearing-level separation.

| **Separation (dB)** | **States queried** | **Matched fraction** |
|---|---|---|
| 0 to 2 | 330 | 1.0000 |
| 2 to 4 | 4,265 | 1.0000 |
| 4 to 6 | 13,456 | 0.9994 |
| 6 to 8 | 15,770 | 0.9871 |
| 8 to 10 | 13,655 | 0.9472 |
| 10 to 12 | 10,386 | 0.8565 |
| 12 to 14 | 6,296 | 0.7213 |
| 14 to 16 | 4,320 | 0.5046 |
| 16 to 18 | 2,833 | 0.2926 |
| 18 to 20 | 1,979 | 0.1708 |
| 20 to 22 | 1,492 | 0.0643 |
| 22 to 24 | 781 | 0.0256 |
| 24 to 26 | 477 | 0.0231 |
| 26 to 28 | 260 | 0.0000 |

*Inner-to-outer hair-cell direction at the primary 5 dB criterion. The decline is continuous across bands rather than concentrated at one separation.*

At census scale the limit was a ceiling, not a transition. Across 4,210,288 audiograms, both an outer-only and an inner-only lesion fitted at least 97% of ears up to a mean severity of 18 dB, 43% at 32 dB and none above 44 dB (**Fig. 3b**), the mean severity that complete OHC loss produces in BEZ2018 (19 dB at 500 Hz, 54 dB at 6 kHz). The decline never exceeded what a reach ceiling alone produces.

The multidimensional hearing state was uninformative only at low severity. Below 20 dB mean severity both lesions fitted at least 99% of ears whatever the hearing state, with one exception of 92%. At 25 to 30 dB the share ran from 15% for the most frequency-specific audiograms to 97% for states with moderate high-frequency slope, and at 30 to 35 dB from 0.04% to 77%; adjusted for survey and severity, the spread across hearing states was 0.073 against 0.014 under permutation (4,168,949 ears with a mean severity from 0 to 60 dB and at least one non-zero hearing level, for which the measure is defined).

#### Within the tested biophysical descriptions, these observations state the Law of Cochlear Degeneracy: one kind of cochlear injury can stand in for another only at the same frequency and only up to a point

OHC and IHC damage trade off place by place, so the damage needed at 2 kHz is set by the damage at 2 kHz in the other cell type and not by damage elsewhere along the cochlea; where places are physically coupled the trade reaches no further than a quarter octave. It holds only up to the model-bounded ceiling of isolated OHC loss, 44 dB on average in BEZ2018. The full hearing state is not part of the law: below 20 dB mean severity it says nothing about cell type, and above that it narrows the differential, because the ceiling is lower at low frequencies than at high.

### State-matched lesions retain different cochlear mechanics

To separate physiologically degenerate states, we compared secondary outputs while the audiometric matching stayed unchanged. Using every admissible pole-dominant and *G*_met_-dominant state at the 19.5, 24.0 and 28.5 dB targets (2,037 and 173 distinct states), we compared physiological responses.

Distortion-product otoacoustic emissions separated the two lesion families completely in every tested target-by-frequency condition. The emissions of all pole-dominant states lay below those of every state-matched *G*_met_-dominant state in all 12 target-by-*f*_2_ cells, with absolute standardized separations of 29.5 to 47.5 and median differences at 6 kHz of 9.9, 17.0 and 32.8 dB at the three targets (**Fig. 4a**; **Table 4**).

**Fig. 4.**
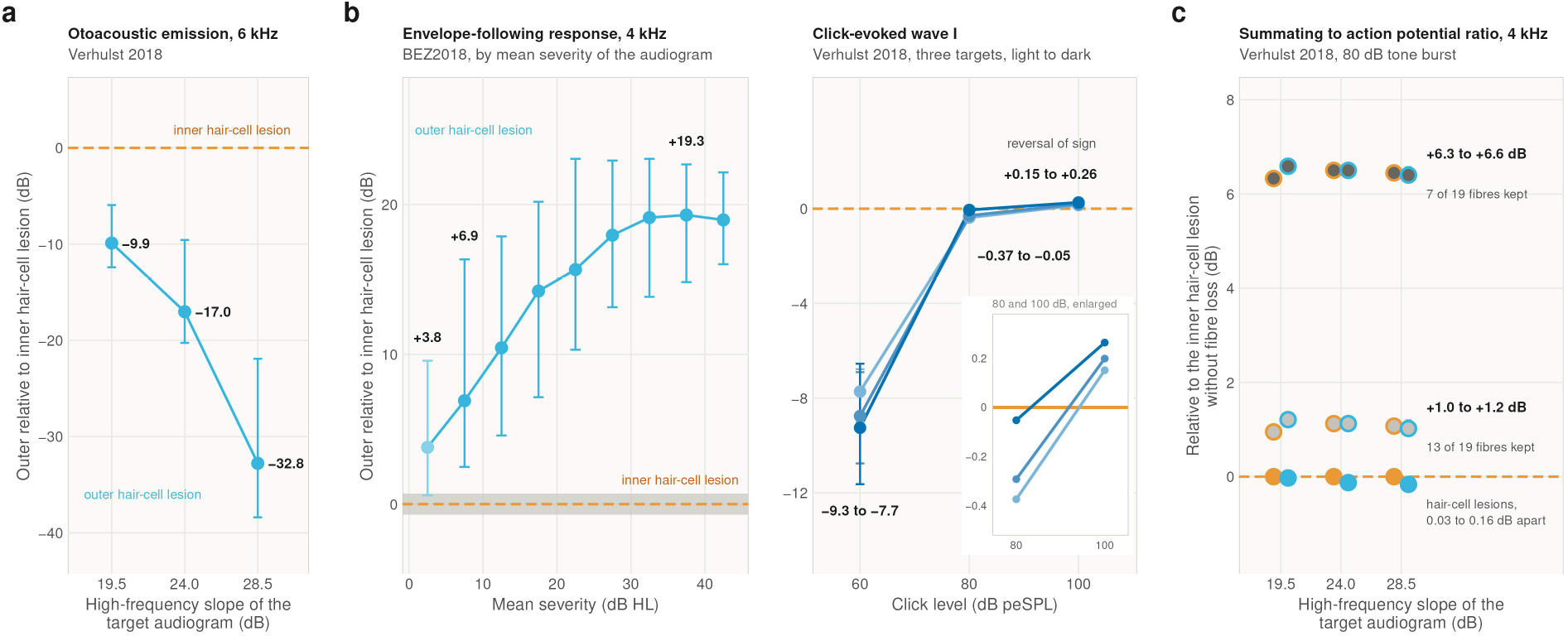
Otoacoustic, envelope-following, auditory-nerve and summating-potential responses of lesions matched on the audiogram. Colours as in Fig. 1, the outer hair-cell lesion blue, the inner hair-cell lesion orange and loss of auditory-nerve fibres dark grey. In a and b the vertical axis is the response of the outer relative to the inner lesion fitting the same audiogram, in decibels; the orange dashed line is the inner lesion. (a) Distortion-product otoacoustic emission at f2 = 6 kHz in Verhulst 2018, gain-loss relative to transduction-loss states at the three target audiograms with high-frequency slope (2,037 and 173 distinct states in all). Points, difference of the medians; bars, the extreme differences between states. Fibre loss leaves the emission unchanged. (b) Left, envelope-following response at 4 kHz in BEZ2018, outer-only relative to inner-only states fitting the same census audiogram, by 5 dB band of mean severity (57,787 pairs of 2,696 audiograms); points, median over pairs; bars, 5th to 95th percentile; grey band, seed-to-seed variation. Every outer-only state lay above every inner-only state in 98.7 to 100% of audiograms from 5 dB HL and in 91.3% below (lighter), short of the 95% rule. Right, click-evoked wave I in Verhulst 2018 for the three targets (light to dark, 19.5, 24.0 and 28.5 dB); gain-loss states are 7.7 to 9.3 dB smaller at 60 dB, within 0.37 dB at 80 dB and 0.15 to 0.26 dB larger at 100 dB at every target, a reversal of sign; inset, 80 and 100 dB with the vertical scale enlarged. (c) Ratio of summating to action potential for a 4 kHz tone burst at 80 dB in Verhulst 2018, relative to the median of transduction-loss states without fibre loss. Filled orange and blue, the two hair-cell lesions, 0.03 to 0.16 dB apart; dark grey, the same states with loss of nerve fibres, keeping 13 (light) or 7 (dark) of 19 fibres per inner hair cell, all high-spontaneous-rate, ringed in the colour of their hair-cell lesion; bars, range over states. Summating and action potentials are model read-outs without an electrode model. Separation is a property of the models and not a clinical recommendation. HL, hearing level.

**Table 4.** Separation of state-matched pole-dominant and transduction-dominant states, by measurement.

| **Measurement** | **Conditions tested** | **Absolute standardized difference** | **Conditions completely separated** |
|---|---|---|---|
| Distortion-product otoacoustic emission | 12 | 29.5 to 47.5 | 12 of 12 |
| Basilar-membrane response | 36 | 16.8 to 115.5 | 36 of 36 |
| Auditory nerve | 36 | 0.15 to 81.8 | 27 of 36 |
| ABR | 27 (16 with a defined difference) | 0.0 to 53.4 | 24 of 27 |
| Envelope-following response | 6 | 15.2 to 40.4 | 6 of 6 |

*Every distinct admissible state at the 19.5, 24.0 and 28.5 dB targets with high-frequency slope (2,037 pole-dominant and 173 transduction-dominant states). A condition is completely separated when the ranges of the two families over all states are disjoint. Eleven ABR conditions had zero within-family variance, so no standardized difference is defined for them. Conditions are target-by-frequency cells for otoacoustic emissions, target-by-frequency-by-level or compression cells for basilar-membrane responses, target-by-level-by-measure cells for auditory-nerve and ABR responses, and target-by-carrier cells for envelope-following responses.*

While mechanistically expected, because *G*_met_ perturbation does not alter the basilar-membrane stage, the result demonstrates that two states with identical audiograms retain divergent cochlear-mechanical outputs.

Basilar-membrane responses showed the same dissociation: all 36 frequency, level and compression cells were completely separated, and at 4 kHz and 40 dB input pole-dominant states were 17, 20 and 25 dB lower at the three targets. At 80 dB the difference became much smaller, so hearing-state equivalence concealed large mechanical differences whose visibility depended on stimulus level.

### State-matched lesions retain different neural physiology

Neural responses differed as well, but depended on stimulus level. Of 36 auditory-nerve cells (three targets, three click levels, four measures), 27 separated pole- and *G*_met_-dominant states completely, as did 24 of 27 auditory brainstem response (ABR) cells (**Fig. 4b**).

Auditory-nerve peak-to-peak output was smaller in pole-dominant states at 60 dB and larger at 100 dB at all three targets, a reversal of sign. ABR amplitude at 100 dB was smaller in pole-dominant states at the 19.5 dB target, overlapped at 24.0 dB (overlap coefficient 0.24) and was larger at 28.5 dB.

Envelope-following responses (EFRs) behaved differently (23). EFR separated the two families completely at both tested carrier frequencies and all three targets, with zero distributional overlap in every carrier-by-target condition and standardized differences of 15.2 to 40.4, largest at 4 kHz for the 28.5-dB target. Pole-dominant states, which gave lower otoacoustic and basilar-membrane responses, gave the larger EFR, so the hidden injury did not scale all outputs together.

A second model extended this across severity. In BEZ2018, 57,787 pairs of outer-only and inner-only states fitting the same audiogram were drawn from up to 300 census audiograms per band across nine bands of mean severity (0 to 45 dB). The EFR at 4 kHz was larger in every outer-only state than in every inner-only state for at least 98.7% of audiograms in each band from 5 dB upward, by a median of 6.9 to 19.3 dB against seed-to-seed variation below 0.7 dB (**Fig. 4b**). Click-evoked responses of high-spontaneous-rate fibres did not separate the pairs at 60 or 80 dB in any band, and at 100 dB only from 25 dB mean severity. Below 5 dB no measurement did.

Loss of auditory-nerve fibres leaves the modelled audiogram unchanged and could accompany either injury. Two fibre-loss profiles lowered wave I and the EFR at 4 kHz by 0.6 to 1.3 dB and by 6.0 to 6.7 dB. The ratio of summating to action potential for a 4 kHz tone burst at 80 dB rose by 1.0 to 1.2 dB and by 6.3 to 6.6 dB and did not differ between the two hair-cell injuries, so it identified fibre loss whichever hair cell was injured (**Fig. 4c**). The EFR at 4 kHz could not tell transduction injury from mechanical injury with the milder fibre loss, whereas wave I to a 60 dB click placed transduction injury 7.7 to 15.2 dB above mechanical injury with or without fibre loss.

Matching the six-frequency audiogram therefore did not make the modelled cochleae physiologically equivalent: which hidden injury remained recoverable depended on the measurement chosen and on how strongly the system was driven.

### From physiological degeneracy to the Cochlear State Differential

#### Applied to one audiogram, the Hidden Map of the Cochlea Library returns its Hearing-State Cochlear Map: the set of simulated cochlear states that reproduce it within 5 dB at all six frequencies, summarised by the lesion categories to which those states belong

It states which cochlear lesions remain possible for that ear and which are excluded. Resolving it requires a Cochlear State Differential: a targeted secondary physiological exam, which narrows the compatible cochlear lesions and leaves the Differential Cochlear States.

#### The audiogram defines the Hearing-State Cochlear Map; an additional physiological measurement can narrow it when it interrogates physiology not already determined by the audiogram

For each differential the models name a measurement that resolves it: for mechanical against transduction injury, otoacoustic emissions or basilar-membrane response; for outer-only against inner-only injury, the EFR at 4 kHz from 5 dB mean severity, and none below it; for fibre loss on either injury, the summating-to-action-potential ratio at 4 kHz (**SI Appendix, Fig. S2**).

This model-derived assignment is the basis for prospective clinical validation, not a standalone clinical guideline.

## Discussion

The central finding of this study is that the cochlea is physiologically degenerate: distinct cellular injuries remain entirely compatible with the same observable auditory function. At the resolution of clinical audiometry, at least 97.3% of 4.2 million audiograms admitted more than one underlying lesion category. The pure-tone audiogram is therefore an accurate description of hearing, but an inherently ambiguous description of the cochlear state that produces it. Because different abnormalities generate the exact same hearing state, the clinical task must shift from selecting a single best-fit lesion to defining the Cochlear State Differential: identifying which lesions remain possible, which are excluded, and which secondary measurement is required to set them apart.

This diagnostic bounding is a clinical necessity for precision otology and therapeutics. In a recent multicentre trial of gene therapy for otoferlin deficiency, all 42 participants had a total loss of hearing function at baseline, yet the number of otoacoustic emissions present before treatment was significantly associated with the therapeutic outcome (24). Two ears with the same audiogram can differ in their physiology and in how they respond to treatment.

The Hidden Map of the Cochlea Library provides a population-scale lookup from measured audiograms to compatible cochlear states. While earlier temporal-bone histopathology, cohort decomposition, and genomic profiling have observed many-to-one mappings in restricted or non-mechanistic contexts (6, 7, 8, 25, 26, 27, 28, 29), this library quantifies the boundaries of this ambiguity. An external six-frequency audiogram can now be compared directly against it to return its compatible lesion categories, without rerunning the forward simulations.

Crucially, the library also identifies what the models cannot explain. In every tested model, isolated OHC and basilar-membrane dysfunction have a finite range and cannot independently generate profound hearing states, consistent with animal data (30, 31). Furthermore, the 0.24% of audiograms that no tested model could reproduce serves as a population benchmark, highlighting a strict target for future models that must incorporate processes not represented here, such as central-neural ones.

Once the Cochlear State Differential is established, resolving it requires selecting the specific secondary measurement that explicitly separates the cellular states remaining in the differential (32). We demonstrated that while some neural responses overlap, targeted EFRs and otoacoustic emissions can completely dissociate the hidden injuries. Recoverability therefore depends on pairing the appropriate secondary measurement and stimulus regime to the specific ambiguity concealed by the audiogram. A Cochlear State Differential resolved this way remains a physiological inference, and prospective validation against independently established cochlear pathology will be required before these states can be read as definitive tissue measurements.

This study has important limitations. Cochlear lesions were inferred from computational models and were not confirmed by direct tissue examination. Model insufficiency means only that no state within that specific model reproduced the audiogram, not that the analogous tissue process is absent clinically. Additionally, the support maps are conditional on the lesion parameterizations tested. For example, our failure to find a match under a bounded scalar perturbation does not prove that no physiologically distinct cochlear state could produce that hearing state. The Verhulst 2018 physiological comparisons are confined to three targets with high-frequency slope and the BEZ2018 comparisons to sampled audiograms, not the whole population library. The two fibre-loss profiles (all medium- and low-spontaneous-rate fibres removed, with 13 or 7 high-spontaneous-rate fibres kept) were chosen by us rather than fitted to human data, and summating and action potentials were computed without an electrode model. The 36.98 dB boundary applies to the valid parameter range of Verhulst 2018; because that model and CARFAC v2 both represent OHC gain, their agreement is convergent rather than independent. The reported ambiguity also depends on the prespecified 5 dB state-match tolerance and on the 0.95 cut-off used to assign lesion categories. Finally, the pure-tone audiogram was the sole patient-level input, meaning undocumented conductive or central hearing components could be misattributed by the models.

Human hearing therefore presents two complementary forms of order: the observable state is strictly constrained and reproducible, yet the cochlear injuries capable of producing it remain non-unique (33). The next frontier in audiology is not determining which lesion best fits an audiogram, but understanding what physiological quantity different injuries preserve in common, how long that preservation can be maintained, and what biological limit causes it to fail.

## Materials and Methods

### Hidden Map of the Cochlea

The analysis comprised 4,210,288 complete six-frequency audiograms (0.5, 1, 2, 3, 4 and 6 kHz) from four population and occupational surveillance cohorts of the Human Hearing Atlas corpus (5): 4,092,824 from the National Institute for Occupational Safety and Health (NIOSH) occupational hearing-conservation archive (1,365,737 workers, 1981 to 2010) (34), 46,184 from the US National Health and Nutrition Examination Survey (NHANES; 1999 to 2020) (35), 27,712 from the US National Health Examination Survey (NHES; 1963 to 1970) (36) and 43,568 from the Korea National Health and Nutrition Examination Survey (KNHANES; 2009 to 2012) (37). Only complete six-frequency records were included, and no frequency was imputed. The ear-examination is the unit of analysis, because each audiogram is interpreted on its own; repeated examinations of one worker were kept as separate observations, and the multi-category fraction stayed between 88.8% and 97.6% in every survey, including the three single-visit population surveys. Four cochlear descriptions were matched: a phenomenological model of five cochlear and middle-ear mechanisms (M0), OHC and basilar-membrane function (Verhulst 2018), combined OHC and IHC function (BEZ2018/Zilany), and cochlear amplification and frequency tuning (CARFAC v2) (20, 38). A model reproduced an audiogram when its bank held at least one state within 5 dB at all six frequencies. A CARFAC v2 state was six OHC health values between 0 and 1, one at the channel of each audiometric frequency, interpolated linearly across channels; 1,000,000 states from a low-discrepancy sequence were each run through the model at all six frequencies, the hearing level being the level at which on-frequency basilar-membrane output reached its healthy value at the reference level. CARFAC v2 represents OHC injury only and therefore enters no lesion-category result. The library of prespecified cochlear states comprised 16 million combined OHC and IHC states (10 million sampling the global joint domain and 2 million each for OHC-only, IHC-only and near-normal regimes), 4,000,014 states of OHC and basilar-membrane mechanics and 2,999,353 states of the five-mechanism model (M0). Each measured audiogram was matched against every state reproducing it within 5 dB at all six frequencies, the step of clinical pure-tone audiometry (21); every compatible state was retained. Lesion categories were assigned to each state from its own parameters, a parameter counting as impaired below 0.95 of its healthy value (sensitivity analyses 0.98, 0.90 and 0.80), and a category was possible for an audiogram when at least one compatible state of that category existed in any BEZ2018 bank. States were indexed by their five Atlas coordinates, and indexed retrieval matched exhaustive search exactly in every bank. BEZ2018 hearing levels were resolved on a 1.05 dB bisection grid, and replicate evaluations with different random seeds differed by a mean of 0.64 dB, well inside the 5 dB criterion. For the 352,949 distinct audiograms with fewer than 30 compatible BEZ2018 states within 2.5 dB, an adaptive stage generated 13,138,048 further states with the same forward model: per-frequency hearing-level maps placed proposals near each audiogram, every hearing level was computed by the same bisection search as in the prespecified banks, and no lesion-category outcome was used to direct sampling. Each compatible state is constructive evidence for its category, and added states can only add categories, so the multi-category fraction can only rise with sampling depth (89.8% with the prespecified banks, 97.3% with both stages), and 97.3% is a lower bound. With the 1,000,000 CARFAC v2 states the library held 37,137,415 states. To locate where interchangeability ends, the fraction of audiograms compatible with both an outer-only and an inner-only state was computed in 2 dB bins of mean six-frequency hearing level and tested with a prespecified statistic, the largest absolute second difference of that curve, against a reach-ceiling null in which support depends only on how many states a bank holds at each severity (1,000 replicates); a transition was to be called real only above the null’s 99th percentile and twice its median. No curve met the criterion (largest statistic 0.11 against 0.585 and 0.93). For audiograms above the point where half had lost interchangeability, 94.6% of those lacking an outer-only state lay beyond what complete OHC loss generates in BEZ2018 (19 dB at 0.5 kHz, 39 dB at 1 kHz, 50 to 54 dB from 2 to 6 kHz), whereas 99.6% of those lacking an inner-only state lay within that family’s reach, so only the outer-only absence marks a reach limit. State ranges, tolerances, sampling rules and classification rules were fixed before analysis.

### Model parameterization and sampling domains

A BEZ2018 state was twelve health values, *C*_OHC_ and *C*_IHC_ at each of the six audiometric places, drawn from a Halton low-discrepancy sequence. The 10 million joint states drew both values independently and uniformly between 0.001 and 1; the OHC-only and IHC-only banks held the uninjured cell type at 1 and drew the other between 0.001 and 1; near-normal states drew both between 0.95 and 1. The hearing level at each frequency was the level of a 50 ms tone (2.5 ms ramps) at which a high-spontaneous-rate fibre (70 spikes/s) at that characteristic frequency, with the model’s human cochlear tuning and a 100 kHz sampling rate, rose 10 spikes/s above its spontaneous rate, found by seven bisection steps between -10 and 125 dB SPL and expressed relative to the state with both values at 1. A Verhulst 2018 state was the pole value at the six places, drawn from a Halton sequence between 0.036, the normal-hearing value, and 0.302, the largest the model admits, and converted to hearing level through the model’s cochlear-gain-loss table; 2,000,000 states covered the whole domain and the rest were concentrated on the observed range of human audiograms and on the two limits of the domain. Transduction injury scaled *G*_met_ by a factor from 1 to 0.6 in 1,601 steps. The Verhulst targets formed two ladders, ten flat audiograms from 2.5 to 30 dB and audiograms with a high-frequency slope from 5 to 35 dB in 5 dB steps. The five-mechanism model (M0) has five fields, OHC gain, endocochlear potential, IHC loss, basilar-membrane tuning and middle-ear transfer, each a constant plus a linear trend along the cochlea; the constants spanned 0 to 55 dB, 0 to 85 mV, 0 to 60 (capped at 55 dB), -0.4 to 0.4 in tuning quality and 0 to 28 dB, and each trend a quarter of that span either way, drawn from a Halton sequence over the whole domain, the observed human range and its boundary. In Verhulst 2018, emissions were read from the model’s ear-canal emission output over the last 250 ms of a 300 ms two-tone stimulus with 5 ms ramps, and EFRs used 300 ms tones fully modulated at 110 Hz; in BEZ2018 the envelope-following stimulus lasted 250 ms and the response was summed over eight characteristic frequencies within one octave of the carrier. Both models ran at 100 kHz.

### Physiological resolution of audiometrically indistinguishable cochlear states

To test whether additional physiological measurements could distinguish cochlear states compatible with the same hearing state, hearing-state-equivalent candidate states were retained rather than reduced to a single best-fitting state. Candidate states were propagated through the corresponding cochlear implementation to generate independent physiological outputs.

Neural outputs in BEZ2018 were obtained for state-matched twins. Unique census audiograms admitting both an OHC-only and an IHC-only state within 5 dB were sampled, up to 300 per 5 dB band of mean severity from 0 to 45 dB, selected at even intervals across each band without reference to lesion category or physiology; up to 10 states of each family were taken per audiogram and every pair agreeing within 5 dB at all six frequencies was kept (2,696 audiograms, 39,632 states, 57,787 pairs). For each state the auditory-nerve mean rate was summed over the six audiometric places. The click response was the peak-to-trough of that sum in the 15 ms after a 100 µs click at 60, 80 and 100 dB for high-spontaneous-rate fibres and at 80 dB for medium- and low-spontaneous-rate fibres; the EFR was its magnitude at 110 Hz for fully modulated tones of 1 and 4 kHz at 70 dB. Every state was evaluated under two random seeds. A measurement separated a band when, in at least 95% of audiograms, every outer-only state lay on one side of every inner-only state.

Mechanical and transduction effects were tested independently in Verhulst 2018. Cochlear mechanical state was varied through the place-resolved pole parameters, whereas IHC transduction was varied through the scalar *G*_met_ parameter. Distortion-product otoacoustic emission output was evaluated at 2f_1_-f_2_ using f_2_/f_1_ = 1.22, primary levels of 65 and 55 dB, and the same stimulus duration for all conditions. Basilar-membrane velocity at the corresponding cochlear place was evaluated separately. Each perturbation changed only the designated physiological parameter while all remaining parameters were held fixed.

The Verhulst 2018 comparisons used every distinct state admissible at the three targets at a pairwise tolerance of 5 dB (636, 714 and 687 pole-dominant and 40, 89 and 44 *G*_met_-dominant states). A condition was completely separated when the ranges of the two families over all states were disjoint. Click responses at 60, 80 and 100 dB, EFRs at 1 and 4 kHz, tone bursts of 1 and 4 kHz (2 ms ramps, 12 ms plateau) and the 10 dB quality factor of the basilar-membrane click response were computed from the published model chain. The summating potential was the plateau shift of the IHC potential summed over place and the action potential the onset peak-to-trough of the summed auditory nerve; neither includes an electrode model. Fibre loss was imposed through the model’s fibre counts per IHC (13, 3 and 3 high-, medium- and low-spontaneous-rate fibres by default; 13, 0, 0 and 7, 0, 0), which the hearing-level criterion does not read.

A physiological measurement was considered informative only when hearing-state-equivalent candidate states that remained overlapping under the audiogram became separable in the additional physiological measurement.

Locality was tested in Verhulst 2018 by raising the pole parameter over half an octave from baselines of 0.08 and 0.12 and solving for a compensating change centred elsewhere; a pair held when the audiogram returned within tolerance. The effect of the full hearing state used the angle between the six-frequency audiogram vector and the flat direction, in 10° bins. The share of ears fitted by both an outer-only and an inner-only state was adjusted for survey and 2 dB severity band, and its spread across bins compared with 200 permutations of bin labels within survey and severity band. The test covered mean severities from 0 to 60 dB; ears outside that range and ears with all six hearing levels at 0 dB, for which the angle is undefined, were not included (41,339 of 4,210,288).

## Ethics and software

No new ethical approval or informed consent was required for this secondary analysis of publicly available, de-identified data. Ethics approval, consent and governance for the original source cohorts are detailed in SI Appendix, Table S2. Analysis pipelines were implemented in Rust, Julia, C++ and Python on national high-performance computing infrastructure (Roihu supercomputer of CSC, IT Center for Science, Finland).

## Data, Materials, and Software Availability

Source datasets analysed in this study remain available from their originating agencies under their respective access conditions, licences and data-use requirements. The dataset-specific provenance and access routes are listed in SI Appendix, Table S1. No source record-level data are redistributed by this study. Aggregate numerical data supporting the reported analyses and display items are provided where permitted by the governing source terms. The code necessary to reproduce the findings reported in this study is available upon reasonable request.

## Data Availability

https://wwwn.cdc.gov/nchs/nhanes/

https://data.cdc.gov/d/c294-dri5

https://doi.org/10.26616/NIOSHSD-1003-2019-0

https://knhanes.kdca.go.kr

## Acknowledgments

This research is based on data from NHES (1963 to 1970) and NHANES (1999 to 2020), Centers for Disease Control and Prevention, National Center for Health Statistics, Hyattsville, MD. We thank the National Center for Health Statistics for curating and openly releasing these data, NIOSH for curating and openly releasing the Occupational Hearing Loss surveillance dataset (SD-1003-2019-0), and the Korea Disease Control and Prevention Agency for the Korea National Health and Nutrition Examination Survey. We gratefully acknowledge CSC for computational resources on the Roihu supercomputer. This work was supported by a grant from The Finnish ORL-HNS Foundation. The funder had no role in study design, data collection, data analysis, interpretation, or the decision to submit for publication.

