## Supplementary material for "Hidden Map of the Human Cochlea and the Law of Cochlear Degeneracy": SI Appendix

**This PDF file includes:**

Figures S1 to S8
Tables S1 to S3

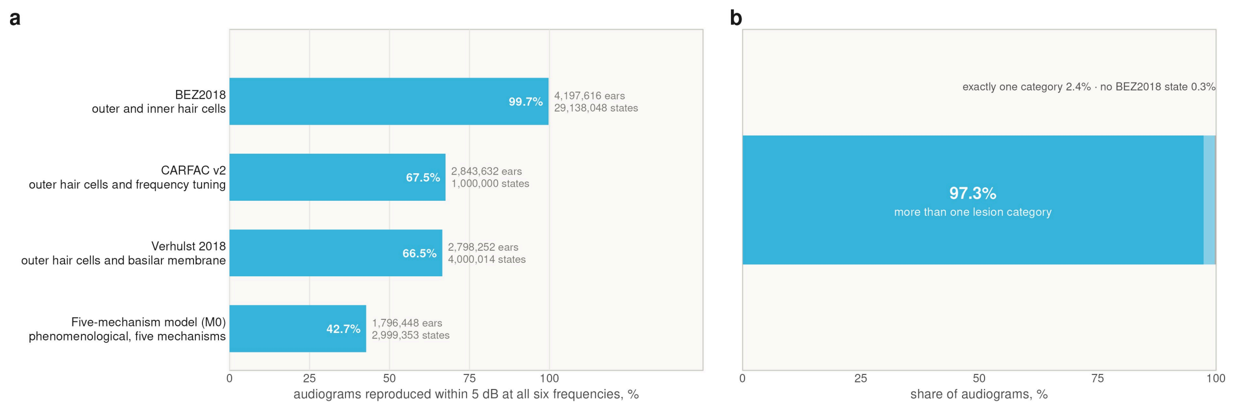


**Fig. S1. Audiogram coverage of four cochlear models and the number of lesion categories compatible with each audiogram.**

(a) Share of 4,210,288 six-frequency audiograms that each of four cochlear models reproduces within 5 dB at all six frequencies, with the ears reproduced and the simulated states of each model. None of the four reproduces 10,270 ears (0.24%).

(b) Lesion categories of the BEZ2018 states compatible with each audiogram at the 5 dB criterion, with more than one category in 97.3% (4,098,154), exactly one in 2.4% (99,462) and none in 0.3% (12,672). The audiograms with none are those BEZ2018 does not reproduce (a), of which 10,270 are reproduced by none of the four models and 2,402 by the five-mechanism model (M0) only. Across the four surveys the share with more than one category ran from 88.8% to 97.6%; added states can only add categories, so each such share is a lower bound.

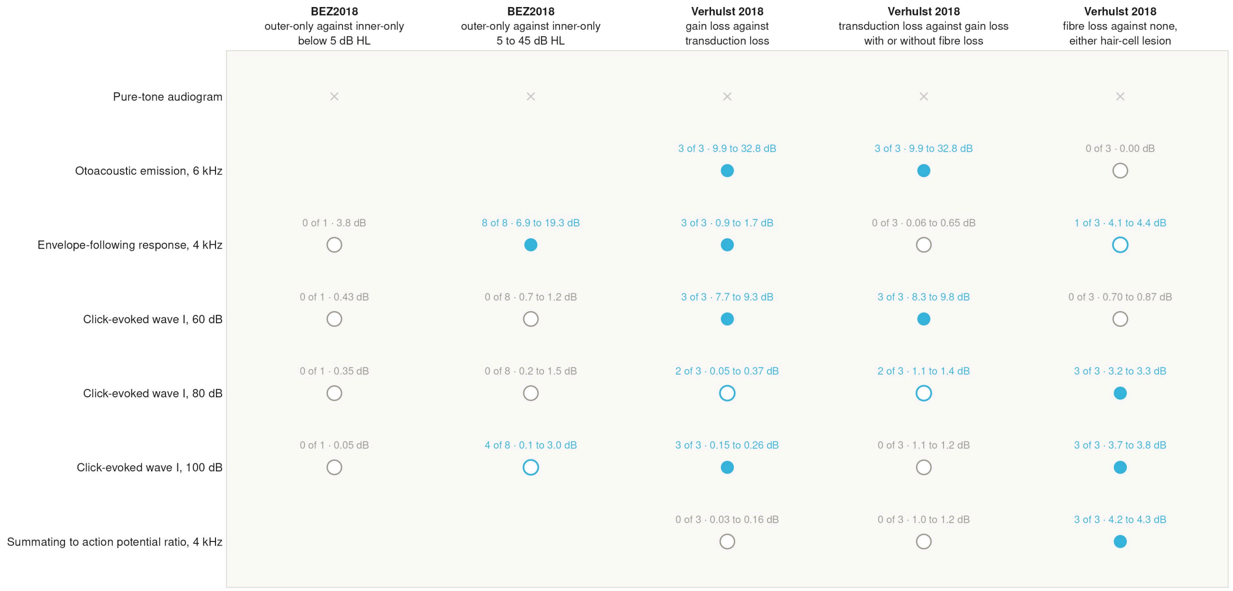


**Fig. S2. Physiological measurements that separate each pair of audiogram-matched lesion types in the models.**

Measurements against differentials, from the model-defined resolution cells (rules in Methods). Strata are the 5 dB bands of mean severity in BEZ2018 (one band below 5 dB HL, eight from 5 to 45 dB HL), with a measurement separating a band when every outer-only state lies on one side of every inner-only state in at least 95% of at least 20 audiograms, and the three targets with high-frequency slope in Verhulst 2018, with a measurement separating a target when the ranges of the two groups over every distinct state are disjoint. A filled circle marks a measurement that separates the two groups of states in every stratum, an open circle of the same colour one that separates them in some strata and a grey open circle one that separates them in none; a cross marks a pair whose states share the audiogram by construction, and a blank cell a measurement not computed in that model. The label above each mark gives the strata separated and the difference between the group medians. Fibre-loss profiles keep 13 or 7 high-spontaneous-rate fibres per inner hair cell and no others. In the fourth column the gain-loss group holds its states without fibre loss and with either profile (wave I at 60 dB places transduction loss 7.7 to 9.3, 8.3 to 9.8 and 13.7 to 15.2 dB above the three taken singly); in the fifth the fibre-loss group holds both profiles and both hair-cell lesions (taken singly, the profiles differ from no fibre loss by 0.6 to 1.3 dB and 6.0 to 6.7 dB, emissions excepted). In BEZ2018 the click-evoked response is the summed response of high-spontaneous-rate fibres; emissions and the summating potential were not computed in that model. Groups can be disjoint at a fraction of a decibel; separation is a property of the models, not a clinical recommendation. HL, hearing level.

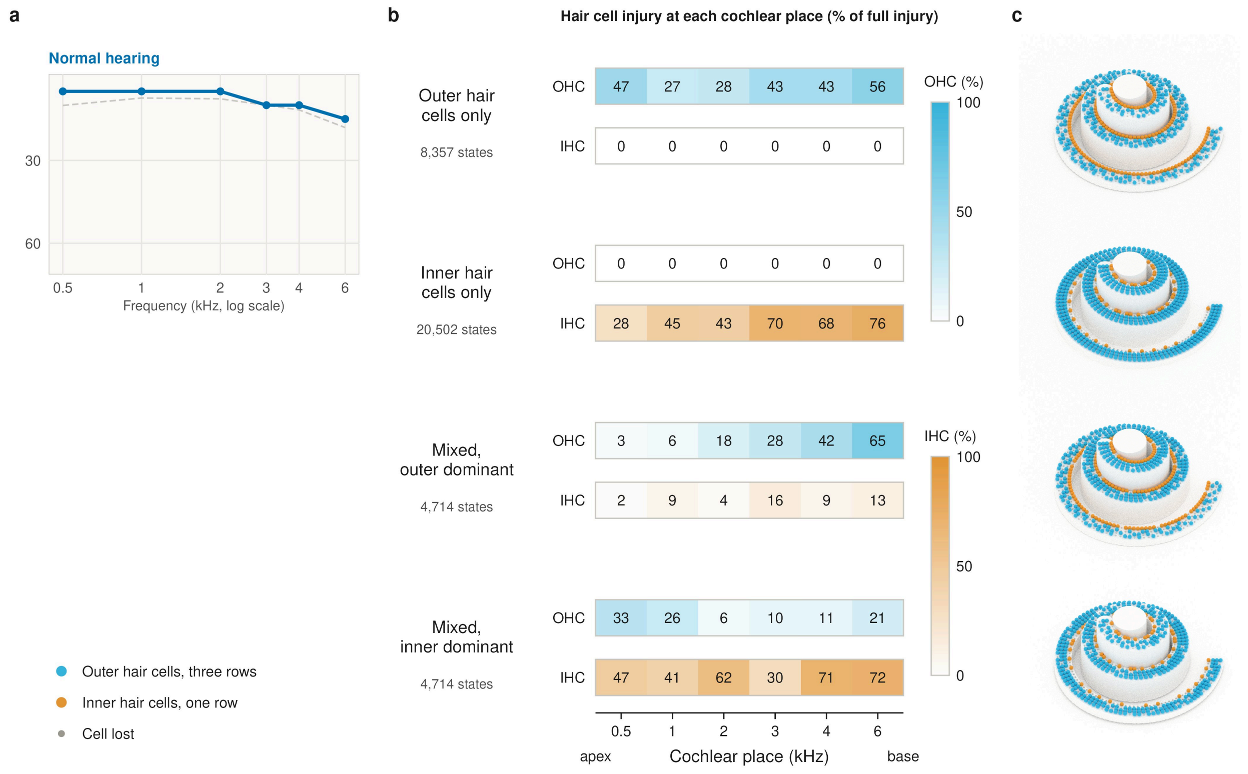


**Fig. S3. Hair-cell injury profiles of simulated cochlear states that reproduce the normal-hearing reference audiogram.**

a, One six-frequency audiogram (hearing state; solid line) against the population median (dashed line). b, Simulated cochlear states (BEZ2018) from its Hearing-State Cochlear Map, each reproducing the audiogram within 5 dB at every frequency: fractional injury of outer hair cells (OHC) and inner hair cells (IHC) at each cochlear place, as a percentage, with the number of library states of that lesion category that reproduce the audiogram. c, The same states drawn as cochleae from the base (6 kHz, outer end) to the apex (0.5 kHz, centre), with three rows of outer hair cells (blue) and one row of inner hair cells (orange); the proportion of cells drawn as lost at each place equals the fractional injury in b. HL, hearing level.

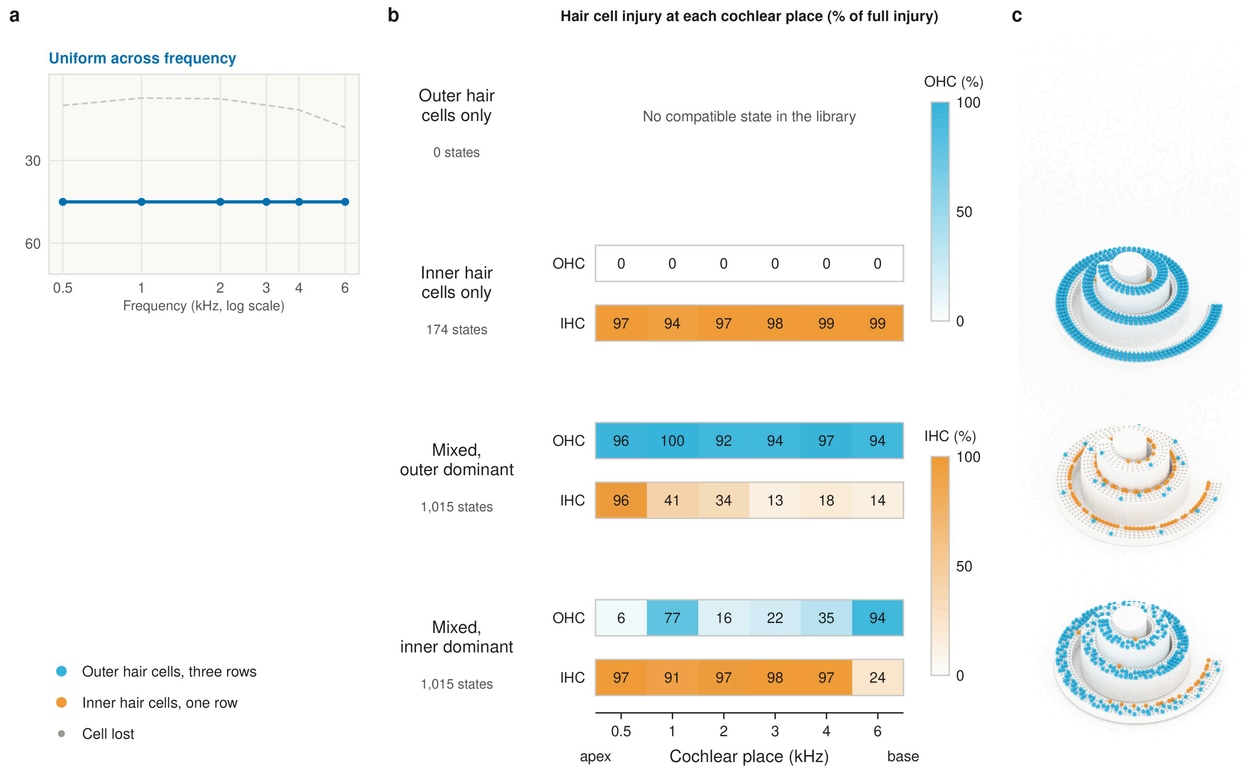


**Fig. S4. Hair-cell injury profiles of simulated cochlear states that reproduce the reference audiogram uniform across frequency.**

a, One six-frequency audiogram (hearing state; solid line) against the population median (dashed line). b, Simulated cochlear states (BEZ2018) from its Hearing-State Cochlear Map, each reproducing the audiogram within 5 dB at every frequency: fractional injury of outer hair cells (OHC) and inner hair cells (IHC) at each cochlear place, as a percentage, with the number of library states of that lesion category that reproduce the audiogram. c, The same states drawn as cochleae from the base (6 kHz, outer end) to the apex (0.5 kHz, centre), with three rows of outer hair cells (blue) and one row of inner hair cells (orange); the proportion of cells drawn as lost at each place equals the fractional injury in b. HL, hearing level.

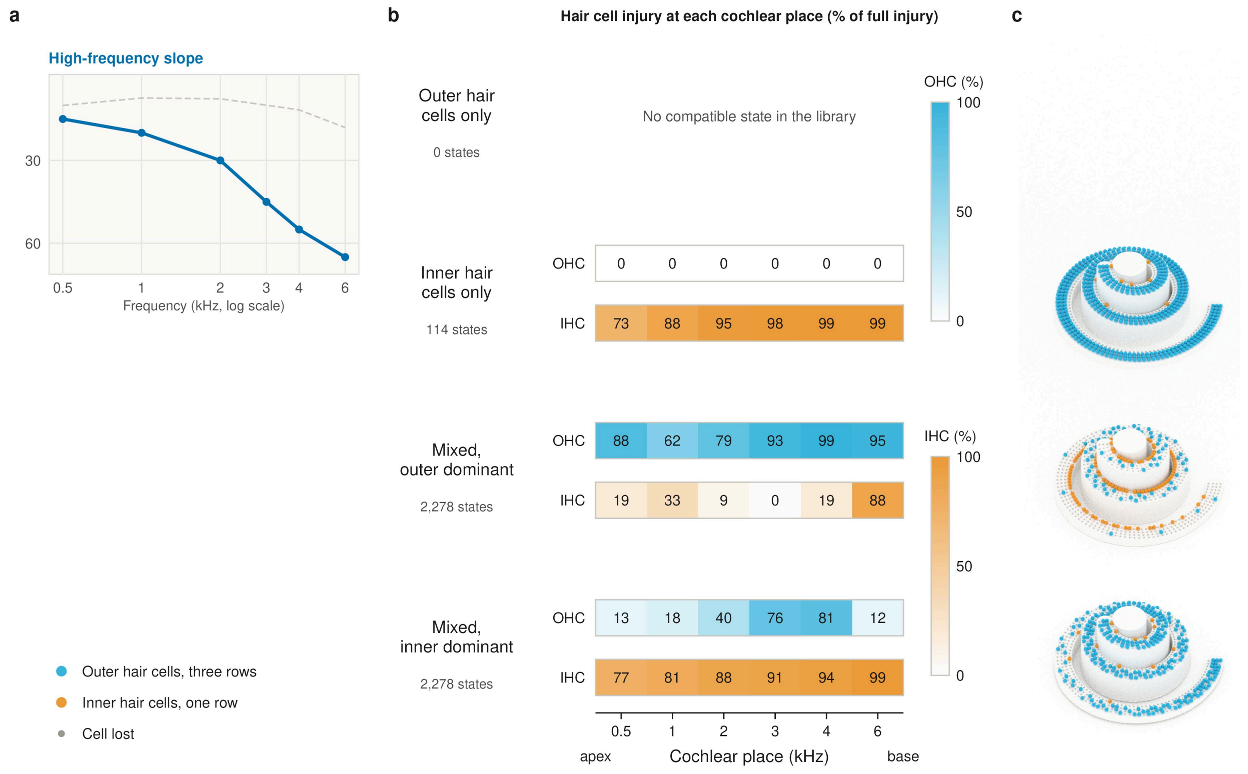


**Fig. S5. Hair-cell injury profiles of simulated cochlear states that reproduce the reference audiogram with a high-frequency slope.**

a, One six-frequency audiogram (hearing state; solid line) against the population median (dashed line). b, Simulated cochlear states (BEZ2018) from its Hearing-State Cochlear Map, each reproducing the audiogram within 5 dB at every frequency: fractional injury of outer hair cells (OHC) and inner hair cells (IHC) at each cochlear place, as a percentage, with the number of library states of that lesion category that reproduce the audiogram. c, The same states drawn as cochleae from the base (6 kHz, outer end) to the apex (0.5 kHz, centre), with three rows of outer hair cells (blue) and one row of inner hair cells (orange); the proportion of cells drawn as lost at each place equals the fractional injury in b. HL, hearing level.

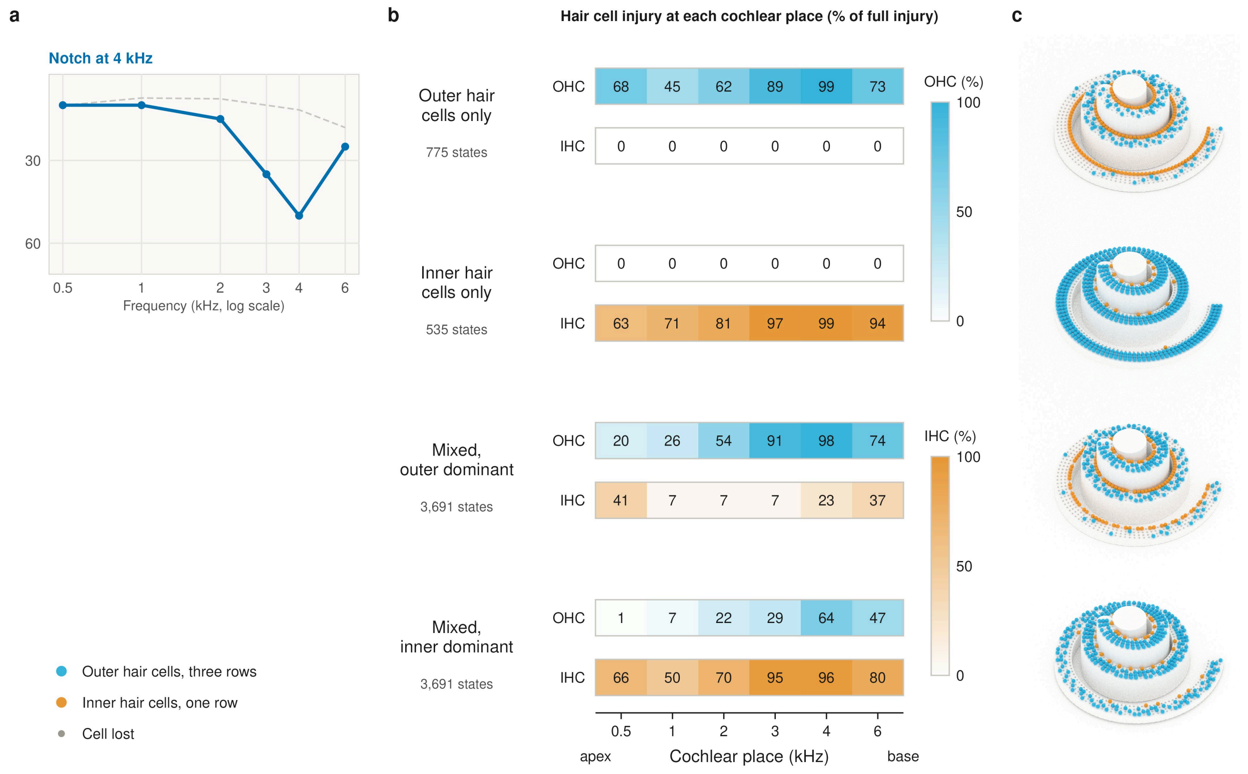


**Fig. S6. Hair-cell injury profiles of simulated cochlear states that reproduce the reference audiogram with a notch at 4 kHz.**

a, One six-frequency audiogram (hearing state; solid line) against the population median (dashed line). b, Simulated cochlear states (BEZ2018) from its Hearing-State Cochlear Map, each reproducing the audiogram within 5 dB at every frequency: fractional injury of outer hair cells (OHC) and inner hair cells (IHC) at each cochlear place, as a percentage, with the number of library states of that lesion category that reproduce the audiogram. c, The same states drawn as cochleae from the base (6 kHz, outer end) to the apex (0.5 kHz, centre), with three rows of outer hair cells (blue) and one row of inner hair cells (orange); the proportion of cells drawn as lost at each place equals the fractional injury in b. HL, hearing level.

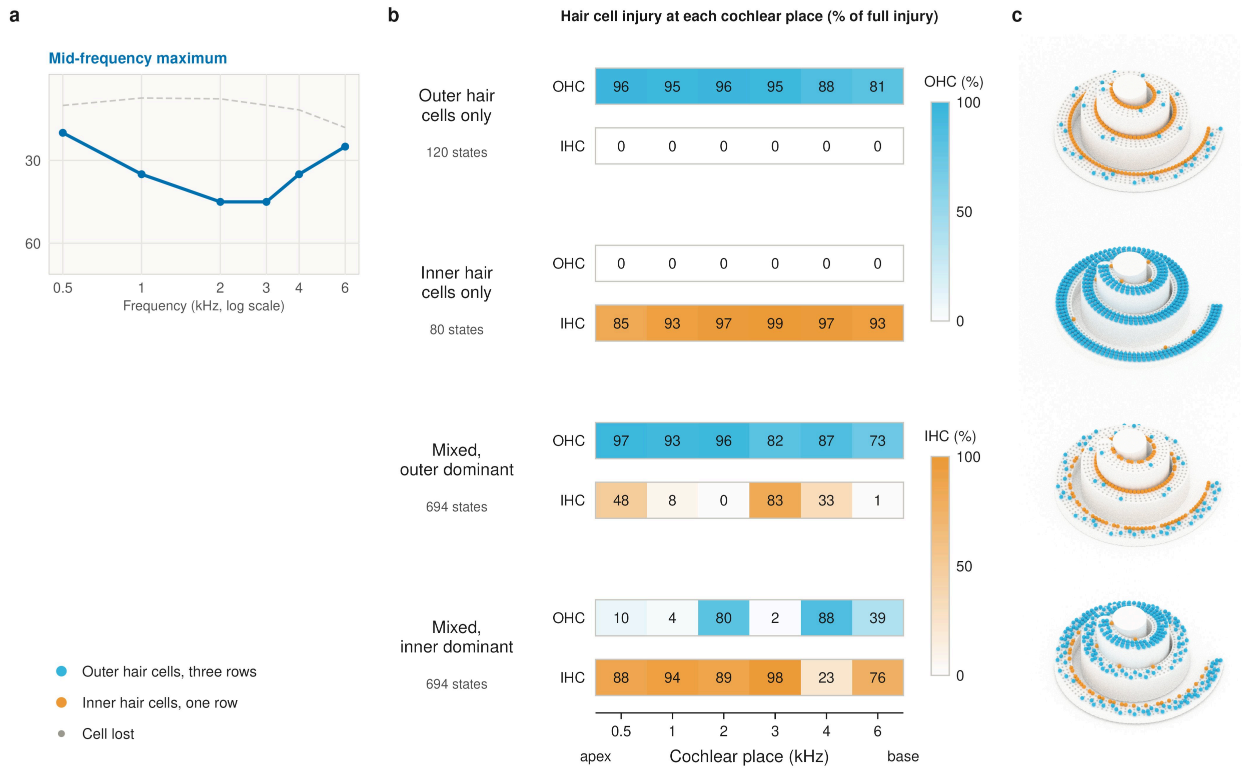


**Fig. S7. Hair-cell injury profiles of simulated cochlear states that reproduce the reference audiogram with a mid-frequency maximum.**

a, One six-frequency audiogram (hearing state; solid line) against the population median (dashed line). b, Simulated cochlear states (BEZ2018) from its Hearing-State Cochlear Map, each reproducing the audiogram within 5 dB at every frequency: fractional injury of outer hair cells (OHC) and inner hair cells (IHC) at each cochlear place, as a percentage, with the number of library states of that lesion category that reproduce the audiogram. c, The same states drawn as cochleae from the base (6 kHz, outer end) to the apex (0.5 kHz, centre), with three rows of outer hair cells (blue) and one row of inner hair cells (orange); the proportion of cells drawn as lost at each place equals the fractional injury in b. HL, hearing level.

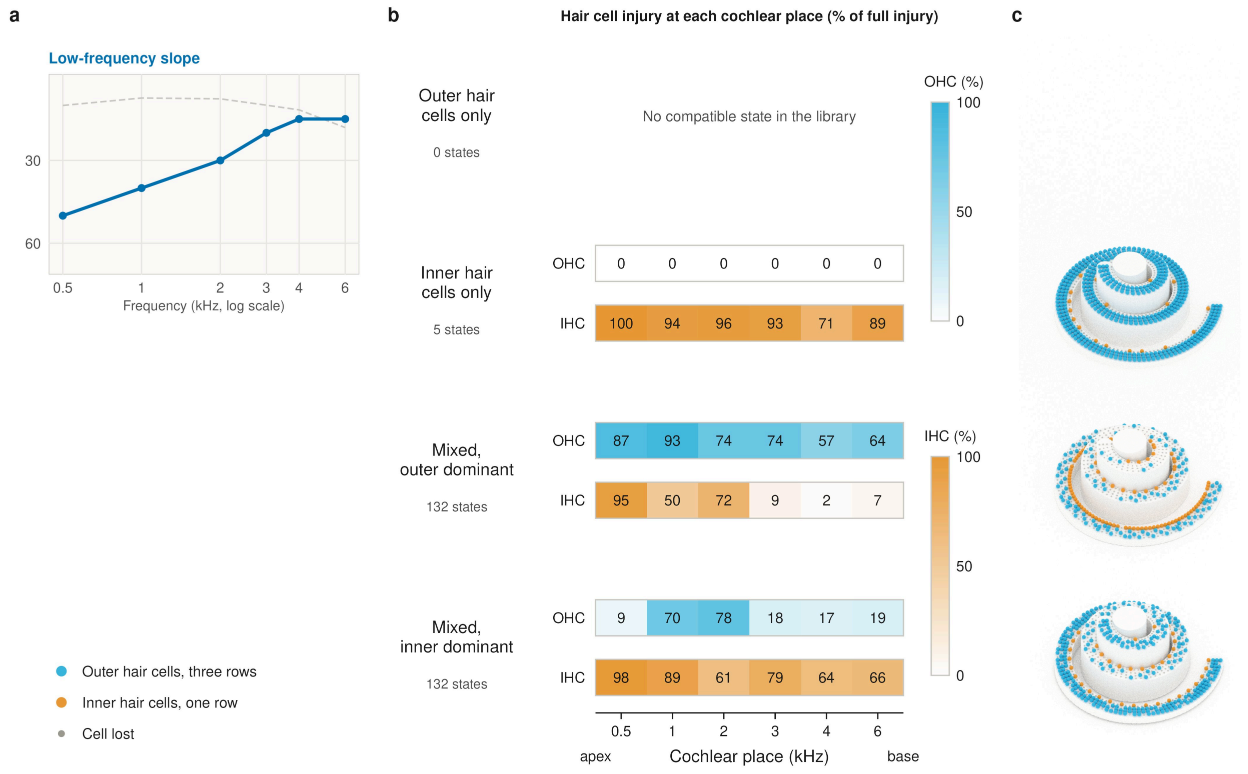


**Fig. S8. Hair-cell injury profiles of simulated cochlear states that reproduce the reference audiogram with a low-frequency slope.**

a, One six-frequency audiogram (hearing state; solid line) against the population median (dashed line). b, Simulated cochlear states (BEZ2018) from its Hearing-State Cochlear Map, each reproducing the audiogram within 5 dB at every frequency: fractional injury of outer hair cells (OHC) and inner hair cells (IHC) at each cochlear place, as a percentage, with the number of library states of that lesion category that reproduce the audiogram. c, The same states drawn as cochleae from the base (6 kHz, outer end) to the apex (0.5 kHz, centre), with three rows of outer hair cells (blue) and one row of inner hair cells (orange); the proportion of cells drawn as lost at each place equals the fractional injury in b. HL, hearing level.

### Table S1. Source datasets, ear-examinations analysed and access routes

| Dataset | Ear-examinations | Source and access | Terms |
|---|---:|---|---|
| NIOSH occupational hearing-conservation archive, United States, 1981 to 2010 | 4,092,824 | Trends in Worker Hearing Loss by Industry Sector, 1981-2010, NIOSH dataset SD-1003-2019-0, https://doi.org/10.26616/NIOSHSD-1003-2019-0; CDC open-data portal, https://data.cdc.gov/d/c294-dri5; download without registration | Public access; Open Database License as listed in the United States federal data catalogue |
| US NHANES, 1999 to 2020 | 46,184 | National Center for Health Statistics, https://wwwn.cdc.gov/nchs/nhanes/; download without registration | Public use; National Center for Health Statistics data-use restrictions |
| US National Health Examination Survey, 1963 to 1970 | 27,712 | National Center for Health Statistics, https://wwwn.cdc.gov/nchs/nhanes/; download without registration | Public use; National Center for Health Statistics data-use restrictions |
| Korea NHANES, 2009 to 2012 | 43,568 | Korea Disease Control and Prevention Agency, https://knhanes.kdca.go.kr; registration required | Agency data-use agreement |
| Total | 4,210,288 | | |

*All data are secondary and de-identified. Ear-examinations are those recorded at all of 0.5, 1, 2, 3, 4 and 6 kHz.*

**Table S2. Ethics approval, consent and governance of the source cohorts**

| Dataset or cohort | Ethics or approval | Consent or governance |
|---|---|---|
| NHES II (1963-1965) | Historical US Public Health Service and NCHS federal survey oversight (pre-Common Rule cohort) | Public-use, de-identified NCHS survey release |
| NHES III (1966-1970) | Historical US Public Health Service and NCHS federal survey oversight (pre-Common Rule cohort) | Public-use, de-identified NCHS survey release |
| NHANES 1999-2004 | NCHS Protocol #98-12 | Informed consent; parental permission and child assent documented |
| NHANES 2005-2010 | NCHS Protocol #2005-06 | Informed consent framework documented |
| NHANES 2011-2016 | NCHS Protocol #2011-17 | Adult consent plus child assent and parental permission |
| NHANES 2017-March 2020 | Protocol #2011-17, then #2018-01 from 26 October 2017 | Written consent; parental consent and assent for minors |
| KNHANES 2009-2012 | KCDC IRB 2009-01CON-03-2C, 2010-02CON-21-C, 2011-02CON-06-C, 2012-01EXP-01-2C | Consent and governance documented by KDCA |
| NIOSH OHL, 1981-2010 | NIOSH IRB determined the programme was research not involving human subjects because audiograms were de-identified | Occupational and regulatory audiograms; de-identified before transfer to NIOSH |

#### Table S3. Lesion-category compatibility by impairment cut-off

| Impairment cut-off (fraction of healthy value) | Audiograms | More than one category (%) | Exactly one category (%) | No category (%) |
|---|---:|---:|---:|---:|
| 0.98 | 4,210,288 | 97.34 | 2.36 | 0.30 |
| 0.95 (primary) | 4,210,288 | 97.34 | 2.36 | 0.30 |
| 0.90 | 4,210,288 | 97.34 | 2.36 | 0.30 |
| 0.80 | 4,210,288 | 97.33 | 2.37 | 0.30 |
| 0.95, mean hearing level 20 dB or more | 959,027 | 89.06 | 10.34 | 0.59 |

*A parameter counts as impaired below the stated fraction of its healthy value. Compatibility is at the 5 dB criterion against the combined BEZ2018 library; added states can only add categories, so each multi-category value is a lower bound.*
